# Multi-Omics Signatures of Alcohol Use Disorder in the Dorsal and Ventral Striatum

**DOI:** 10.1101/2021.10.04.21264523

**Authors:** Lea Zillich, Eric Poisel, Josef Frank, Jerome C. Foo, Marion M. Friske, Fabian Streit, Lea Sirignano, Stefanie Heilmann-Heimbach, André Heimbach, Per Hoffmann, Franziska Degenhardt, Anita C. Hansson, Georgy Bakalkin, Markus M. Nöthen, Marcella Rietschel, Rainer Spanagel, Stephanie H. Witt

## Abstract

Alcohol Use Disorder (AUD) is a major contributor to global mortality and morbidity. Postmortem human brain tissue enables the investigation of molecular mechanisms of AUD in the neurocircuitry of addiction. We aimed to identify differentially expressed (DE) genes in the ventral and dorsal striatum between individuals with AUD and controls, and to integrate the results with findings from genome- and epigenome-wide association studies (GWAS/EWAS) to identify functionally relevant molecular mechanisms of AUD. DNA-methylation and gene expression (RNA-seq) data was generated from postmortem brain samples of 48 individuals with AUD and 51 controls from the ventral striatum (VS) and the dorsal striatal regions caudate nucleus (CN) and putamen (PUT). We identified DE genes using DESeq2, performed gene-set enrichment analysis (GSEA), and tested enrichment of DE genes in results of GWASs using MAGMA. Weighted correlation network analysis (WGCNA) was performed for DNA-methylation and gene expression data and gene overlap was tested. Differential gene expression was observed in the dorsal (FDR<0.05), but not the ventral striatum of AUD cases. In the VS, DE genes at FDR<0.25 were overrepresented in a recent GWAS of problematic alcohol use. The *ARHGEF15* gene was upregulated in all three brain regions. GSEA in CN and VS pointed towards cell-structure associated GO-terms and in PUT towards immune pathways. The WGCNA modules most strongly associated with AUD showed strong enrichment for immune response and inflammation pathways. Our integrated analysis of multi-omics data sets provides further evidence for the importance of immune-and inflammation-related processes in AUD.

## Introduction

Alcohol Use Disorder (AUD) is a major contributor to the global disease burden, with a prevalence of ∼17% among 12-month alcohol users in the US^1, 2^ and an estimated heritability of 49%^3^. Knowledge about the molecular mechanisms can foster understanding of causes and promote prevention. Recent genome-wide association studies (GWASs) have identified 29 genetic loci associated with Problematic Alcohol Use (PAU), a proxy of AUD^4^. While GWASs identify increasing numbers of disease-associated loci, the functional interpretation of many of these findings remains inconclusive. Analyzing the transcriptome can extend the understanding of the molecular mechanisms underlying AUD, by identifying associated gene expression patterns. Findings can in turn be integrated with results from GWASs and epigenome-wide association studies (EWASs) to identify the pathomechanisms underlying disease.

Processes in the central nervous system are considered to play a major role in the etiology of addiction, and the transition from chronic alcohol consumption to AUD^5^. Therefore, it is of particular interest to examine molecular changes associated with addiction in brain tissue. So far, only few studies have been conducted in postmortem human brain tissue to identify transcriptional changes associated with AUD^6-8^. These studies mainly focused on the prefrontal cortex (PFC) one important part of the neurocircuitry of addiction^9, 10^. The first transcriptome-wide study in the PFC found DE genes implicated in neuronal processes, such as myelination, neurogenesis, and neural diseases, as well as cellular processes, such as cell adhesion and apoptosis^11^. In Brodmann Area 9 downregulation of calcium signaling pathways has been observed in individuals with AUD compared to controls^12^. In the same study, a weighted gene co-expression analysis (WGCNA) pointed towards modules associated with AUD case/control status, which were enriched for nicotine and opioid signaling, as well as immune processes. Another study in the PFC (Brodmann Area 8) showed that co-expression networks associated with lifetime alcohol consumption were enriched for GWAS signals of alcohol dependence^6^.

Despite the importance of striatal regions in addiction processes, genome-wide human omics studies of these brain regions are still missing. The striatum is divided into the ventral striatum (VS), consisting of the nucleus accumbens and olfactory tubercle; and the dorsal striatum, comprising the caudate nucleus (CN) and putamen (PUT)^13^. The nucleus accumbens is involved in mediating motivational processes such as aversion and reward, which play a significant role in the development and maintenance of substance use disorders (SUD)^13^. In addition to regulating motor function, the CN and PUT are involved in cognitive processes relevant for addiction, such as executive functioning and cognitive control, reinforcement learning and habit formation^14^. Analyses of omics data from striatal regions could complement the knowledge on global molecular changes in the neurocircuitry of addiction in AUD.

In a recent EWAS of AUD in postmortem brain tissue, we identified differentially methylated CpG-sites and regions in the ventral and dorsal striatum^15^. Previous studies have shown the utility of integrating epigenetic and transcriptomic data in postmortem brain tissue of SUDs using weighted correlation network analysis (WGCNA)^16^. WGCNA clusters genes or CpG-sites into co-expressed or co-methylated modules based on correlation matrices. By relating modules to each other, WGCNA can be used for data integration, providing more insights than descriptive overlap. For example, whereas a descriptive comparison of histone H3 lysine 4 trimethylation (H3K4me3) and mRNA expression in individuals with AUD and cocaine use disorder revealed no consistent overlap between H3K4me3 trimethylation and gene expression^17^, a network analysis identified overlapping modules pointing towards co-expressed genes associated with H3K4me3 trimethylation^6^. Modules associated with AUD were enriched for CNS functions, such as synaptic transmission and regulation of neurogenesis^6^. WGCNA has also been used for integrating epigenetic and transcriptomic data and investigating their association with opioid use disorder (OUD) in postmortem human brain, identifying immune-related transcriptional regulation to be enriched in co-expressed and co-methylated modules^18^.

The aim of the present study was to investigate differential gene expression associated with AUD status in the ventral and dorsal striatum, relate these to GWAS findings, and to integrate the findings with DNA-methylation data using a network approach (WGCNA) in order to identify functionally relevant molecular mechanisms in AUD.

## Materials and Methods

### Samples

Postmortem human brain tissue from CN, PUT and VS of a total of 48 individuals with AUD and 51 control individuals (68% male) was obtained from the New South Wales Tissue Resource Centre at the University of Sydney. The Ethics Committee II of the University of Heidelberg approved the study (reference number 2021-681). After quality control (QC), the total sample sizes for each brain region were N_CN_ = 71, N_PUT =_ 77 and N_VS_ = 63. Phenotypic information was assessed by next-of-kin interviews. Inclusion criteria for this study were: age>18 years, Western European Ancestry, no history of severe psychiatric or neurodevelopmental disorders, or SUDs other than AUD and nicotine use disorder or smoking. AUD was defined as meeting DSM-IV criteria for alcohol dependence and consuming 80g of alcohol a day or more (control group: <20g/day). Descriptive information can be found in Table 1 and Supplementary Table S1.

**Table 1.** Descriptive statistics of demographic data.

| Characteristic | Cases | Controls | *p* |
| --- | --- | --- | --- |
| N | 48 | 51 |  |
| Age, years | 55.58 (10.62) | 57 (10.64) | 0.51 |
| Sex (M/F) | 31/17 | 37/14 |  |
| pH-value | 6.53 (0.26) | 6.65 (0.25) | 0.026* |
| PMI (hours) | 37.07 (15.79) | 30.7 (15.57) | 0.047* |
| Blood Alcohol level (N) | 7 | 0 |  |
| Blood Alcohol Level (g/100ml) | 0.21 (0.21) |  |  |
| Smoking (yes/%) | 32 (66.7%) | 12 (23.5%) | <0.001* |
| Samples per Brain Region |  |  |  |
| Caudate Nucleus | 36 | 37 |  |
| Putamen | 35 | 42 |  |
| Ventral Striatum | 31 | 32 |  |

*Data are presented as count (n/n; n (%)) or mean ( ± SD), PMI: post-mortem interval, pH: pH-value of the brain, p: p-value of t-Test/Chi-squared test comparing cases and controls.*
*\*significant difference between cases and controls*

### RNA extraction and -sequencing

RNA was extracted from frozen tissue according to the manufacturer’s protocol using the Qiagen RNeasy microKit (Qiagen, Hilden, Germany). The RNA Integrity Number (RIN) of all samples was determined using a TapeStation 4200 (Agilent, Santa Clara, CA). RIN values of 273 samples were larger than 5.5, for which libraries were prepared using the TruSeq Stranded Total RNA Library Prep Kit (Illumina, San Diego, CA). RNA sequencing was performed on the NovaSeq 6000 (Illumina) at the Life & Brain Center in Bonn, Germany with read lengths of 2×100bp and a sequencing depth of 62.5 M read pairs per sample on average. Technical replicates were sequenced for all but four samples.

### DNA extraction and Methylation Profiling

DNA extraction, methylation profiling, and QC was performed as described in Zillich et al.^15^. In brief, DNA was extracted using the DNeasy extraction kit (Qiagen, Hilden, Germany); the Illumina HumanMethylation EPIC BeadChip and the Illumina HiScan array scanning system (Illumina, San Diego, CA) were used to determine DNA-methylation levels. We used an updated and customized version of the CPACOR pipeline to extract beta values from raw intensities^19^. Criteria for the removal of samples and probes can be found in Zillich et al.^15^. In the present analyses, DNA methylation data was included from all subjects from whom gene expression data was available after QC.

### Statistical Analyses

All analyses apart from QC and read mapping were performed using R version 3.6.1^20^. An overview of the analysis workflow can be found in Figure 1. The Benjamini-Hochberg (FDR)^21^ procedure was used to correct for multiple testing. Differentially expressed genes were considered statistically significant at FDR<0.05. All downstream analyses were performed using genes significantly differentially expressed at FDR<0.25.

**Figure 1.**
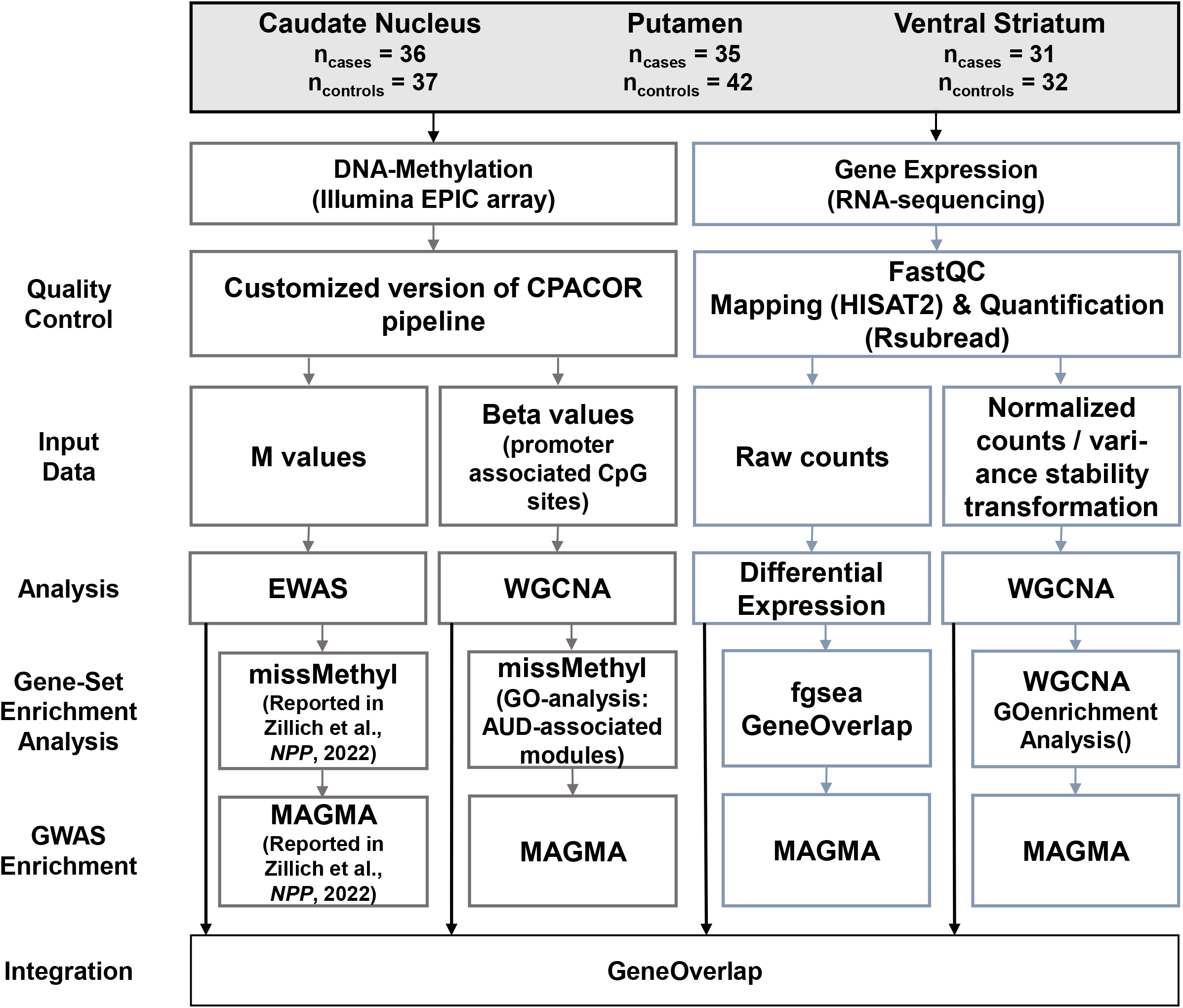
Analysis workflow of the present study.

#### Mapping and Quantification

Sequencing quality was determined using FastQC^22^ and 24 samples (11 cases and 13 controls) were excluded due to insufficient sequencing quality (e.g. strong overrepresentation of sequences, GC distribution). Raw reads were mapped to the human genome (hg38) using HISAT2 (v.2.1.0)^23^. Quantification was performed with the featureCounts function of the Rsubread package (v.2.0.1)^24^, with hg38 annotation.

#### Differential Gene Expression Analysis

Differential gene expression was determined using DESeq2 (v.1.26.0)^25^. Minimal pre-filtering was applied, removing genes with normalized counts <10 for more than two samples. Technical replicates were merged prior to differential expression analysis using the collapseReplicates function as implemented in DESeq2. For the differential gene expression analysis, we included age, sex, RIN, pH-value of the brain, and postmortem interval (PMI) as covariates, because of their known influence on gene expression^26-28^. To assess residual bias after adjustment for covariates, we generated Q-Q plots and calculated genomic inflation factors (Supplementary Figure 1). We further conducted a variance partition analysis using the variancePartition() function of the corresponding R package^29^, which confirmed the covariates. Results of this analysis can be found in Supplementary Figure 2. Results were filtered for differentially expressed (DE) genes with an absolute log2 fold change larger than 0.02. Volcano plots displaying up- and downregulation of genes for each brain region are shown in Supplementary Figure 3.

#### Gene-Set Enrichment Analysis

Gene-set enrichment analysis was performed using the R package fgsea (v.1.12.0)^30^, for which DE genes were ranked according to p-value. Enrichment analysis was performed for Gene-Ontology (GO) terms^31^ and Hallmark gene sets^32^ and the results were adjusted using FDR correction.

#### Cell Type Enrichment Analysis

To identify cell type specific expression signatures, we performed cell type enrichment analysis using DE genes (FDR<0.25) from the three brain regions. As a reference gene set for brain cell types, we used the “top ranked cell type-enriched genes based on human data” as provided by McKenzie, Wang ^33^. These contain the 1000 most enriched genes in a cell type and cover astrocytes, endothelial cells, microglia, neurons, and oligodendrocytes. Using the R package GeneOverlap (v.1.22.0)^34^, we assessed the overlap of AUD-status associated DE genes with markers from the different cell populations. Results were adjusted for multiple testing using the Benjamini-Hochberg method as implemented in GeneOverlap.

#### Differential Methylation Analysis

Effect sizes and p values for CpG sites were used from the EWAS results as presented in the original publication^15^. In brief, the EWAS model was based on methylation M-values as the dependent variable and AUD status as the predictor. As covariates, sex, age, postmortem interval (PMI), pH-value, estimated smoking, standardized neuronal cell count, and the first ten principal components of the EPIC array internal control probes were included.

#### WGCNA

Weighted correlation network analyses (WGCNA, v.1.70-3)^16^ were performed to identify modules of co-expressed genes and co-methylated CpG-sites. We assessed the relationship of these modules with AUD case/control status and tested the overlap between associated modules. WGCNA clusters the input matrix according to a dynamic tree cutting algorithm, using a soft power threshold that approximates the criterion of scale-free topology (R_signed_^2^>0.80). Resulting soft power thresholds for expression networks were 6 for CN, 5 for PUT, and 14 for VS; for methylation networks, all power thresholds were 2.

To identify methylation networks associated with gene expression, beta values from normalized intensities of all samples from which gene expression data were available were filtered for promoter-associated CpG-sites based on the manufacturer’s manifest (Illumina, San Diego, CA). The resulting 105 796 CpG-sites were used as input.

For the RNA-seq data, count matrices were normalized using the DESeq2 function normalizeCounts and variance stability transformation was applied.

Networks were constructed using following settings: minimum module size=30, mergeCutHeight=0.25, maxBlockSize=36 000. In WGCNA, modules are labeled using colors. In the results section modules are labeled according to type of data, brain region, and color assigned in the analysis, e.g. “e-VS-pink” for module “pink” from the WGCNA analysis of gene expression data in the ventral striatum. For each module, its eigengene was calculated and correlated with AUD status. Association of modules with AUD status and covariates is shown in Supplementary Figure 4. For modules associated with AUD status, we performed enrichment analysis using the GOenrichmentAnalysis function implemented in the WGCNA package for expression data and the R package missMethyl (v.1.20.4)^35^ for methylation modules. Further, we extracted hub genes of AUD associated WGCNA expression modules by calculating the product of module membership and gene significance for each gene of a module. Based on this score, the 10% of highest ranking genes were defined as module hub genes. To investigate the biological relevance of hub genes, protein-protein interaction networks were generated using the *Search Tool for the Retrieval of Interacting Genes/Proteins* (STRING, v.11.5)^36^. Graphical representation of gene networks was restricted to high confidence interactions (interaction score threshold 0.7).

#### Expression and Methylation Data Integration

To identify genes both DE and differentially methylated, we analyzed the overlap of DE genes (FDR<0.25) with the results of an EWAS (p<0.001) in the same sample^15^. We prioritized CpG-sites based on their functional relevance in gene expression regulation. Thus, promoter-associated CpG-sites were used in the analysis.

At the module level, gene-set overlap tests were performed using the R package GeneOverlap (v.1.22.0)^34^. Here, Fisher’s exact test is used to identify significant overlap. For each brain region, the overlap of the AUD-associated co-expression and co-methylation modules was tested.

#### GWAS Enrichment Analysis

We analyzed enrichment of DE genes with an FDR<0.25, and genes in AUD-associated WGCNA modules in GWAS summary statistics using Multi-marker Analysis of GenoMic Annotation (MAGMA, v.1.08b)^37^. We performed GWAS enrichment analysis for several SUDs, such as alcohol use disorder and problematic alcohol use^4^, cannabis use disorder^38^, and a recent GWAS comparing individuals with opioid use disorder with unexposed controls^39^. Bonferroni correction (n=4 tests per gene set) of p values was applied to adjust for multiple testing.

## Results

### Differential Gene Expression

Gene expression analysis of postmortem brain tissue from AUD cases and controls revealed DE genes at FDR<0.05 in both dorsal striatal regions. In the caudate nucleus, 49 DE genes were identified at FDR<0.05 (39 up- and 10 downregulated). Tubulin Tyrosine Ligase Like 4 (*TTLL4*, log2FC=0.11, p=2.3*10^−8^) and GATA Binding Protein 2 (*GATA2*, log2FC=-0.27, p=8.6*10^−7^) were the most significantly upregulated and downregulated genes, respectively. Top up- and downregulated genes in the putamen were found to be Transcription Elongation Factor A Like 2 (*TCEAL2*, log2FC=0.09, p=5.8*10^−5^) and Desmin (*DES*, log2FC=-0.86, p=2.6*10^−6^), the latter being the only significant gene after correction for multiple testing. Nine genes were downregulated in both dorsal striatal regions, with *HLA-DOB* having the highest log2FC in both regions. In the ventral striatum, no DE genes were detected at FDR<0.05. The most significant differential gene expression in the ventral striatum was observed for Ankyrin Repeat And Ubiquitin Domain Containing 1 (*ANKUB1*) which was upregulated in AUD cases (log2FC=1.35, p=5.8*10^−5^). In the VS of AUD cases Caseinolytic Mitochondrial Matrix Peptidase Chaperone Subunit B (*CLPB*, log2FC=-0.11, p=5.2*10^−6^) was the most significantly downregulated gene.

None of the DE genes at FDR<0.05 overlapped between multiple brain regions. Therefore, the less conservative significance threshold of FDR<0.25, which was also used for downstream analyses, was applied to compare the overlap of DE genes. At FDR<0.25 the cardiomyopathy associated 5 (*CMYA5*) gene showed an upregulation in both caudate nucleus and putamen. *ARHGEF15* (Rho Guanine Nucleotide Exchange Factor 15) was upregulated in all three brain regions at FDR<0.25. The Top 5 DE genes from each brain region are listed in Table 2; complete summary statistics are listed in Supplementary Table S2 (CN), S3 (PUT), and S4 (VS). Overlap between DE genes in the different brain regions is shown in Figure 2A.

**Table 2.**

| Entrez Gene ID | Gene Name | baseMean | log2(FC) | lfcSE | Stat | P-Value | FDR |
| --- | --- | --- | --- | --- | --- | --- | --- |
| Caudate Nucleus |  |  |  |  |  |  |  |
| 9654 | *TTLL4* | 1125.89 | 0.11 | 0.02 | 5.59 | $2.33 \times 10^{-8}$ | 0.0005 |
| 2624 | *GATA2* | 51.17 | -0.27 | 0.05 | -4.92 | $8.58 \times 10^{-7}$ | 0.0091 |
| 25904 | *CNOT10* | 695.68 | 0.06 | 0.01 | 4.84 | $1.27 \times 10^{-6}$ | 0.0091 |
| 222256 | *CDHR3* | 1483.68 | 0.19 | 0.04 | 4.75 | $1.99 \times 10^{-6}$ | 0.0106 |
| 375611 | *SLC26A5* | 63.80 | 0.28 | 0.06 | 4.62 | $3.81 \times 10^{-6}$ | 0.0163 |
| Putamen |  |  |  |  |  |  |  |
| 1674 | *DES* | 22.06 | -0.86 | 0.18 | -4.70 | $2.64 \times 10^{-6}$ | 0.0486 |
| 2050 | *EPHB4* | 118.04 | -0.19 | 0.05 | -4.19 | $2.78 \times 10^{-5}$ | 0.0939 |
| 9144 | *SYNGR2* | 499.84 | -0.20 | 0.05 | -4.19 | $2.76 \times 10^{-5}$ | 0.0939 |
| 55741 | *EDEM2* | 348.56 | -0.07 | 0.02 | -4.23 | $2.30 \times 10^{-5}$ | 0.0939 |
| 84245 | *MRI1* | 662.19 | -0.13 | 0.03 | -4.13 | $3.57 \times 10^{-5}$ | 0.0939 |
| Ventral Striatum |  |  |  |  |  |  |  |
| 81570 | *CLPB* | 1188.85 | -0.11 | 0.02 | -4.56 | $5.16 \times 10^{-6}$ | 0.0653 |
| 22899 | *ARHGEF15* | 54.23 | -0.26 | 0.06 | -4.49 | $7.04 \times 10^{-6}$ | 0.0653 |
| 55584 | *CHRNA9* | 5.58 | -1.17 | 0.26 | -4.44 | $9.14 \times 10^{-6}$ | 0.0653 |
| 100463488 | *MTRNR2L10* | 4.91 | -2.07 | 0.50 | -4.16 | $3.23 \times 10^{-5}$ | 0.1730 |
| 389161 | *ANKUB1* | 30.26 | 1.35 | 0.34 | 4.02 | $5.79 \times 10^{-5}$ | 0.2480 |

*Top 5 differentially expressed genes in caudate nucleus, putamen and ventral striatum.*

**Figure 2.**
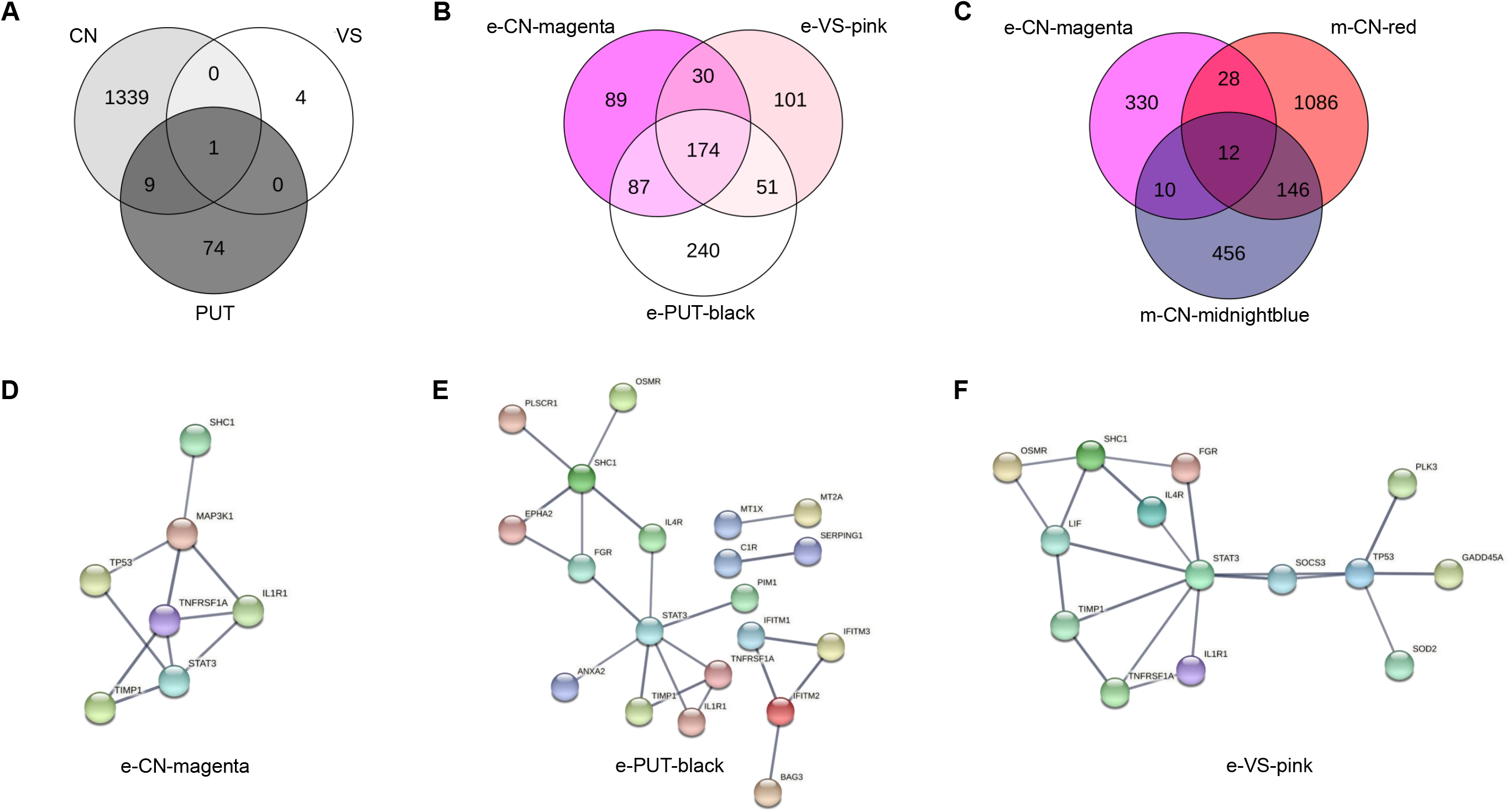
Venn Diagrams of gene overlap of A) DE genes at FDR < 0.25 in caudate nucleus (CN), putamen (PUT), and ventral striatum (VS), B) genes forming WGCNA expression-modules showing the strongest association with AUD status for CN, PUT, and VS, C) genes forming WGCNA expression-module “e-CN-magenta” and those forming the methylation-modules “m-CN-red” and “m-CN-midnightblue”. Network plots depicting the WGCNA gene expression modules showing the strongest association with AUD: D) module magenta from caudate nucleus, E) module black from putamen and F) module pink from ventral striatum.

### Gene-set Enrichment Analysis

Pathway analysis using a pre-ranked enrichment analysis revealed significant enrichment of dorsal striatum DE genes for several GO terms and Hallmark gene-sets. Genes in the CN were found to be related to cilia- and microtubule-associated GO-terms, while none of the Hallmark gene-sets was significantly enriched. GO-term and Hallmark gene-set analysis in PUT samples showed enrichment for immune processes, such as “acute inflammatory response to antigenic stimuli” (p_FDR_=0.006) and “adaptive immune response” (p_FDR_ =0.006). In the VS the most significantly enriched GO-terms were also related to cilia and microtubules, as well as antigen processing. All GO-terms and Hallmark gene-set with FDR <0.10 are listed in Supplementary Tables S5 (CN), S6 (PUT), and S7 (VS).

### Cell Type Enrichment Analysis

In the CN, upregulated DE genes were significantly enriched for astrocytic markers (p_FDR_=7*10^−6^), whereas an enrichment for endothelial cell marker genes was detected among downregulated genes (p_FDR_ =2*10^−7^). No significant cell-type enrichment of DE genes was found in the putamen and the ventral striatum. GeneOverlap heatmap visualizations for the three brain regions are displayed in Supplementary Figure 5.

### WGCNA

#### Expression

In the CN, 21 modules with a median size of 352 genes (range: 64-7 259) were identified. Module “e-CN-magenta”, consisting of 328 genes, showed the strongest positive association with AUD status (r=0.42, p=2.89*10^−4^). In the PUT, of the 25 modules (median size 249 genes, range: 33-5 381) identified, module “e-PUT-black” was most strongly correlated with AUD with a positive direction of effect (r=0.41, p=2.31*10^−4^). For expression data from the ventral striatum, 16 modules with a median size of 429 genes (range: 35-9 708) were identified; module “e-VS-pink” had the strongest positive association with AUD (r=0.41, p=0.009). Interestingly, in a GO-term analysis the three AUD associated modules were all enriched for immune processes, such as “defense response” and “inflammation response”. Gene network representation of hub genes in modules “e-CN-magenta”, “e-VS-pink”, and “e-PUT-black” revealed the signal transducer and activator of transcription 3 (*STAT3*) gene as a conserved hub node in all three brain regions (Figure 2D-2F). There was also a wide overlap of the genes in the three modules: 174 (22.54%) were partially shared between all three modules corresponding to the three brain regions, while another 21.76% were shared between at least two modules (Figure 2B). A gene network analysis of the 174 shared genes between regions identified *STAT3, TP53, ICAM1, MYC*, and *NFKBIA* as the top 5 hub nodes of the network. A visualization of the network is depicted in Figure 3.

**Figure 3.**
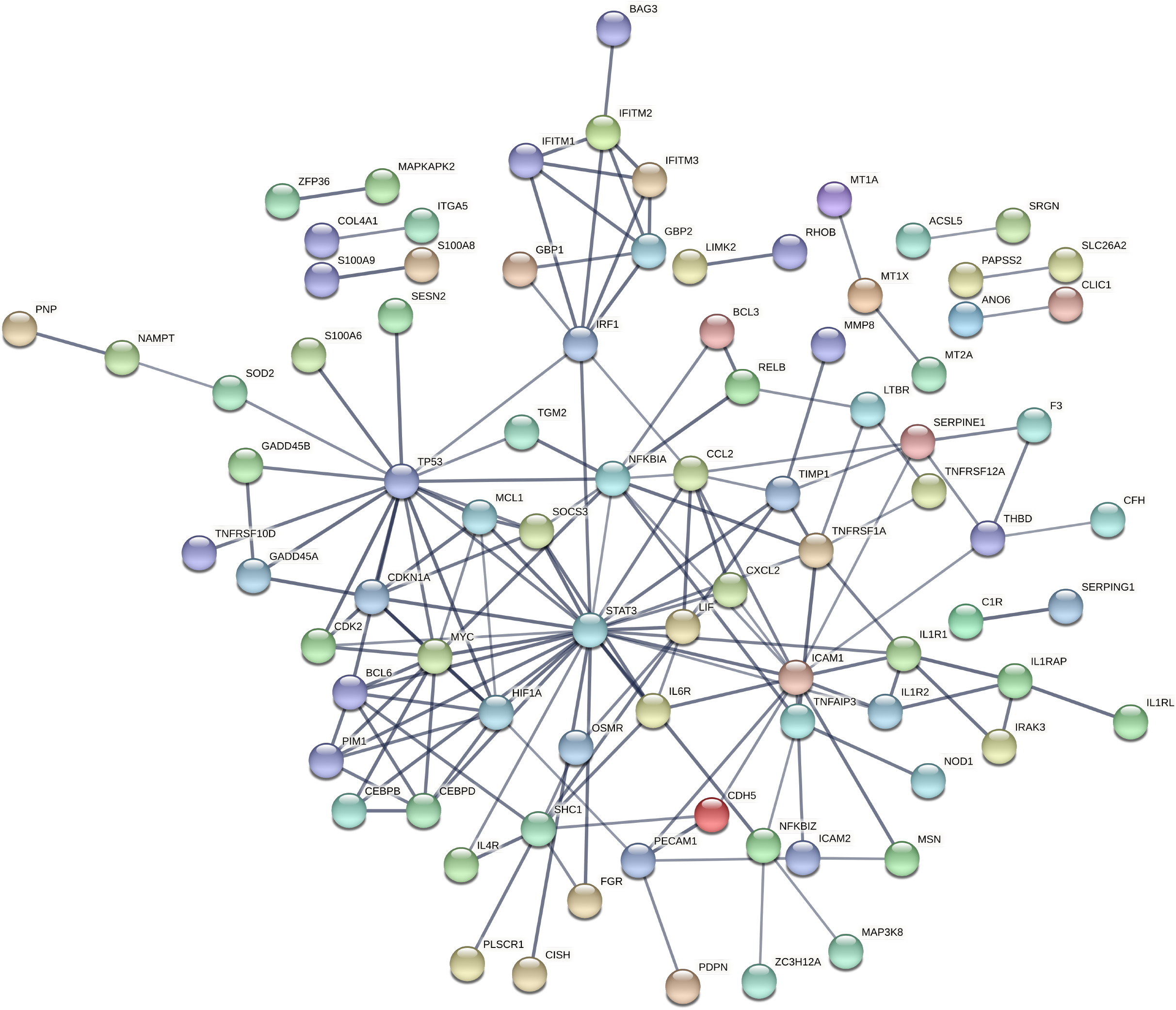
Network plot of genes, co-expressed between the WGCNA modules e-CN-magenta, e-PUT-black and e-VS-pink.

#### Methylation

In the CN, WGCNA resulted in 36 modules with a median size of 346 CpG-sites (range: 66-41 423). Module “m-CN-red”, consisting of 2 117 CpG-sites, showed the strongest association with AUD case control status (r=-0.27, p=0.021). This module was most highly enriched for the biological processes “cell activation” (p=1.52*10^−5^) and “leukocyte activation” (p=2.09*10^−5^). In PUT 177 modules were identified (median size=57 CpG-sites, range: 30-42 248). Module “m-PUT-plum” consisted of 70 CpG-sites and was significantly associated with AUD case/control status (r=-0.29, p=0.023) and enriched for the biological processes “positive regulation of I-κB kinase/NF-κB signaling” (p=0.002) and “regulation of I-κB kinase/NF-κB signaling” (p=0.005). WGCNA in the VS methylation data resulted in 85 modules (median size=178 CpG-sites, range: 35-30 370). The module with the strongest association with AUD was “m-VS-lavender” (r=-0.29, p=0.023), which consisted of 117 CpG-sites and was enriched for the molecular function “natural killer cell lectin-like receptor binding” (p=3.43*10^−4^) and the biological process “susceptibility to natural killer cell mediated cytotoxicity” (p=3.65*10^−4^). The top 10 enriched GO-terms for all AUD-associated modules can be found in Supplementary Tables S8-S10.

### Expression and Methylation Data Integration

In the CN, 12 genes showed both differential methylation and differential gene expression. DE statistics, EWAS summary statistics and functional annotation for these genes are provided in Supplementary Table S11. No overlap was observed in the VS and PUT. At the module-level, co-expression module “e-CN-magenta” showed significant overlap with the methylation modules “m-CN-red” (p=0.003) and “m-CN-midnightblue” (p=0.014) (Figure 2C), while expression module “e-CN-purple” did not show significant overlap with the methylation modules in CN. Of the 3 AUD-associated expression modules in the VS, only “e-VS-salmon” showed significant overlap with the methylation module “m-VS-turquoise” (p=0.003), but not “m-VS-lavender”. No overlap was observed for gene expression and DNA-methylation in PUT.

### GWAS Enrichment Analysis of DE Genes and WGCNA modules

In the VS, but not in the dorsal striatum, we observed enrichment of DE genes in the GWAS signal of PAU (p=0.045). In the putamen DE genes were enriched for GWAS signal from a study comparing individuals with OUD to unexposed controls (p=0.025). None of the DE genes in any of the brain regions showed enrichment for signals from a GWAS of Cannabis Use Disorder or AUD.

From the WGCNA modules showing the strongest association with AUD, only module e-VS-pink showed significant enrichment for GWAS signals of CUD (p=0.043). None of the findings remained statistically significant after multiple testing correction. Results from the respective analyses are depicted in Supplementary Figure 6 and enrichment p values as well as the number of overlapping genes are displayed in Supplementary Table S12.

## Discussion

In the present study, we identified DE genes, co-expression networks, and pathways associated with AUD in the dorsal and ventral striatum. The results were integrated with DNA-methylation data and results from GWASs of SUDs.

We discovered that one gene (*ARHGEF15*) was consistently upregulated in all investigated brain regions of AUD cases compared to controls. *ARHGEF15* encodes a specific guanine nucleotide exchange factor for the activation of Ras homolog family member A (RhoA), a GTPase, which has been linked to higher blood pressure and hypertension over the Rho/ROCK signaling cascade^40^. It is postulated that the Rho Guanine Nucleotide Exchange Factor 15 negatively regulates excitatory synapse development by suppressing the synapse-promoting activity of EPHB2^41^. EPHB2 deficiency has been linked to depression-like behaviors and memory impairments in animal studies^42^. In line with this, genetic variation within *ARHGEF15* has been associated with hematocrit, red blood cell count, and hemoglobin concentration^43^, but also with psychiatric traits, such as neuroticism and worries^44^ as well as bipolar disorder^45^.

Among the genes that were downregulated in both dorsal striatal regions, *HLA-DOB* displayed the highest fold change. HLAs of the Major Histocompatibility Class II are an essential part of the acquired immune system presenting antigens to T-lymphocytes (for review: Howell, Carter ^46^). The most significantly downregulated gene in the VS is *CLPB*, a mitochondrial chaperone, which has been associated with progressive brain atrophy^47^ and with the cellular response to alcohol-induced stress^48^. In a recent GWAS, *CLPB* was associated with the amount of alcohol consumed on a typical day (p=9.67*10^−5^, N=116 163)^49^.

DE genes in the ventral striatum were enriched for GWAS signals of PAU, but not AUD. This could be a result of the larger sample size of the PAU GWAS, but also point towards differences in genetic variation as responsible for differential expression.

Our results from the pathway and network analyses further underline immune-related effects of chronic alcohol exposure; the pathway and network modules most strongly associated with AUD case-control status were also enriched for immune system and inflammation processes. This was observed for all three brain regions, and both in expression and methylation data, providing further evidence for the important role of immune processes in AUD.

Gene networks derived from WGCNA hub genes similarly revealed genes related to inflammatory processes as strongly connected network nodes. Here, *STAT3* represents a conserved network hub node in all three brain regions. *STAT3* is a member of the JAK/STAT pathway and acts as a transcription factor upon activation by cytokines, hormones and growth factors^50^. Interestingly, a recent study assessing expression signatures of alcohol withdrawal in rats discovered a very similar gene network in the hippocampus with *STAT3* as a hub node surrounded by a network of downstream targets^51^. The authors also discovered increased levels of STAT3 and its neuroinflammation-related target genes in postmortem brain tissue of subjects with AUD. Activation of the *STAT3* gene network was found to be primarily restricted to astrocytes. This supports the results of the cell type enrichment analyses, where enrichment of astrocytic expression signatures was detected for upregulated DE genes in the CN.

These results strongly reflect the well-described effect of chronic alcohol exposure on different aspects of the innate and acquired immune systems^52^. Chronic alcohol exposure accelerates the inflammatory response and reduces anti-inflammatory cytokines^52^. An activated immune response in response to chronic alcohol exposure has been shown on the cell level^53^, as well as on the transcription^53^, and protein levels^54, 55^. In a previous EWAS, we found strong enrichment of immune processes in differentially methylated CpG-sites associated with alcohol withdrawal^56^. Neuroinflammation has been repeatedly associated with AUD and both the glutamate excitotoxicity and the production of acetaldehyde, key processes in AUD metabolism, have been suggested to produce an inflammatory response in the brain^57^. On a phenotypic level, there is also widespread overlap between symptoms of inflammation and of SUDs, such as anhedonia, depression, and decreased cognitive functioning^58^. In addition, in candidate gene studies in postmortem human PFC, hippocampus, and orbitofrontal cortex, increased mRNA levels of *HMGB1*, which encodes a proinflammatory cytokine and toll-like receptor genes have been associated with alcohol consumption in AUD cases, providing evidence for chronic neuroinflammation in response to alcohol^59-61^. Notably, there is an overlap of findings not only on the single-gene level but also on the level of pathways and networks/modules. This overlap underlines that alcohol consumption has common biological effects in different brain regions, i.e., most prominently, effects on immune and inflammation processes.

Several limitations apply to our study. First, we cannot distinguish between effects being a consequence of chronic alcohol consumption or addiction. Second, although we corrected for PMI, which can influence tissue quality as a confounding factor, it cannot be ruled out that other characteristics not easily accounted for, such as cause of death, or blood alcohol level for which the majority of individuals have missing data, influenced gene expression. Third, although the sample size is comparatively large for postmortem brain studies in the addiction field, the small number of differentially expressed genes is likely attributable to limited power. Lastly, analyzing bulk tissue does not adequately reflect the diversity of cell types across different brain regions and future studies on single-cell level are needed to investigate cell-type specific transcriptional changes associated with AUD.

It has to be noted that besides DNA methylation, epigenetic mechanisms such as histone and chromatin modifications, or microRNA expression profiles can influence gene expression and are especially important in addiction research^62^. Future studies should therefore expand the epigenetic profiling of AUD to include these mechanisms.

In summary, the present study provides further evidence from multi-omics data sets for the importance of immune-and inflammation-related processes in AUD. Notably, drugs that reduce neuroinflammation to reduce drinking, such as phosphodiesterases, may be promising approaches for novel treatment options for AUD. Recently published randomized controlled trials suggest that a phosphodiesterase inhibitor reduces heavy drinking whereas an antibiotic compound was not effective^63, 64^. A deeper understanding of the underlying mechanisms will enhance the discovery of drug targets and drive forward the development of precision medicine within in this field.

## Data Availability

Raw data are available on request. Summary statistics are presented in the Supplementary Materials.

## Acknowledgments

The study was supported by the German Federal Ministry of Education and Research (BMBF), “A systems-medicine approach towards distinct and shared resilience and pathological mechanisms of substance use disorders” (01ZX01909), “Towards Targeted Oxytocin Treatment in Alcohol Addiction” (031L0190). ERA-NET: Psi-Alc (01EW1908), and the Deutsche Forschungsgemeinschaft (DFG) – Project-ID 402170461 – TRR 265^65^.

We thank Elisabeth Röbel and Claudia Schäfer-Arnold for technical assistance.

Tissues were received from the New South Wales Brain Tissue Resource Centre at the University of Sydney which is supported by the University of Sydney. Research reported in this publication was supported by the National Institute of Alcohol Abuse and Alcoholism of the National Institutes of Health under Award Number R28AA012725. The content is solely the responsibility of the authors and does not represent the official views of the National Institutes of Health.

## Supplementary Information

Supplementary information is available at MP’s website.

## Data and Code Availability

Raw data and analysis code are available from the corresponding author upon reasonable request.

