## Supplementary figures and images for "Multi-Omics Signatures of Alcohol Use Disorder in the Dorsal and Ventral Striatum"

### Supplementary Figure 1

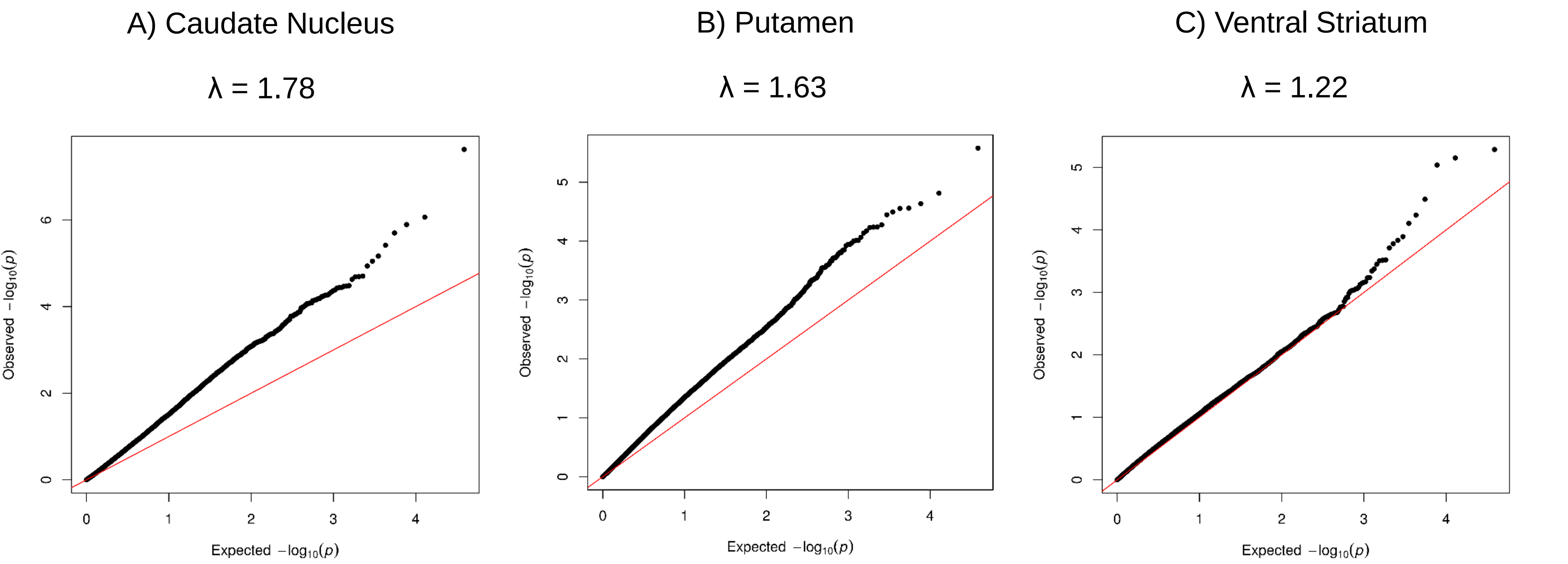

### Supplementary Figure 2

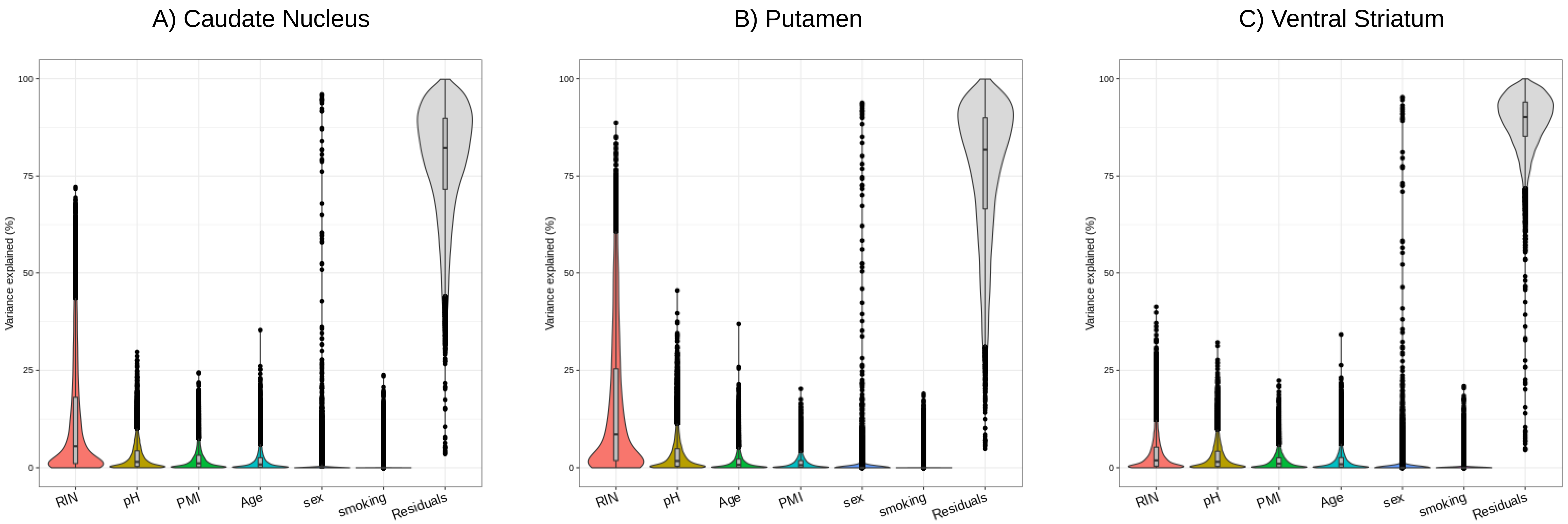

### Supplementary Figure 3

A) Caudate Nucleus

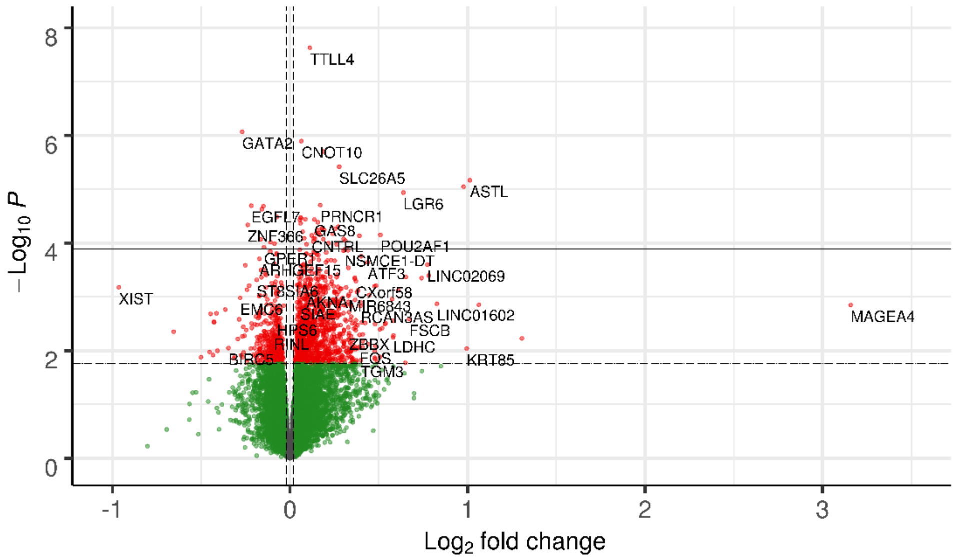


B) Putamen

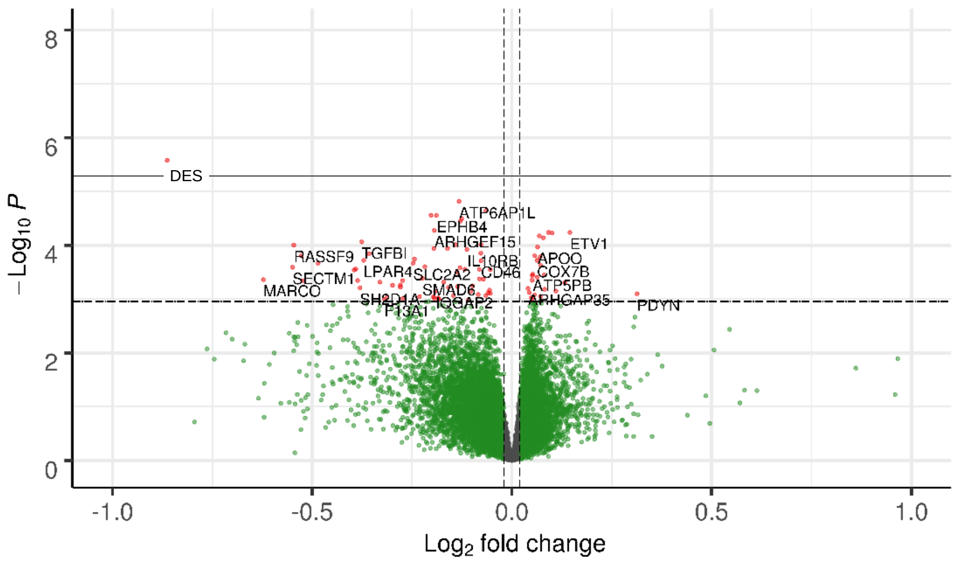


C) Ventral Striatum

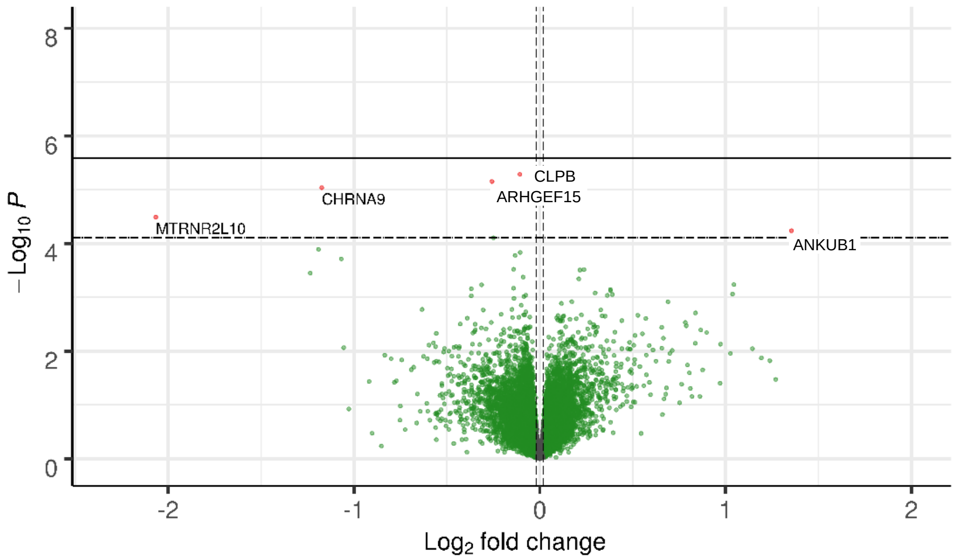

### Supplementary Figure 4

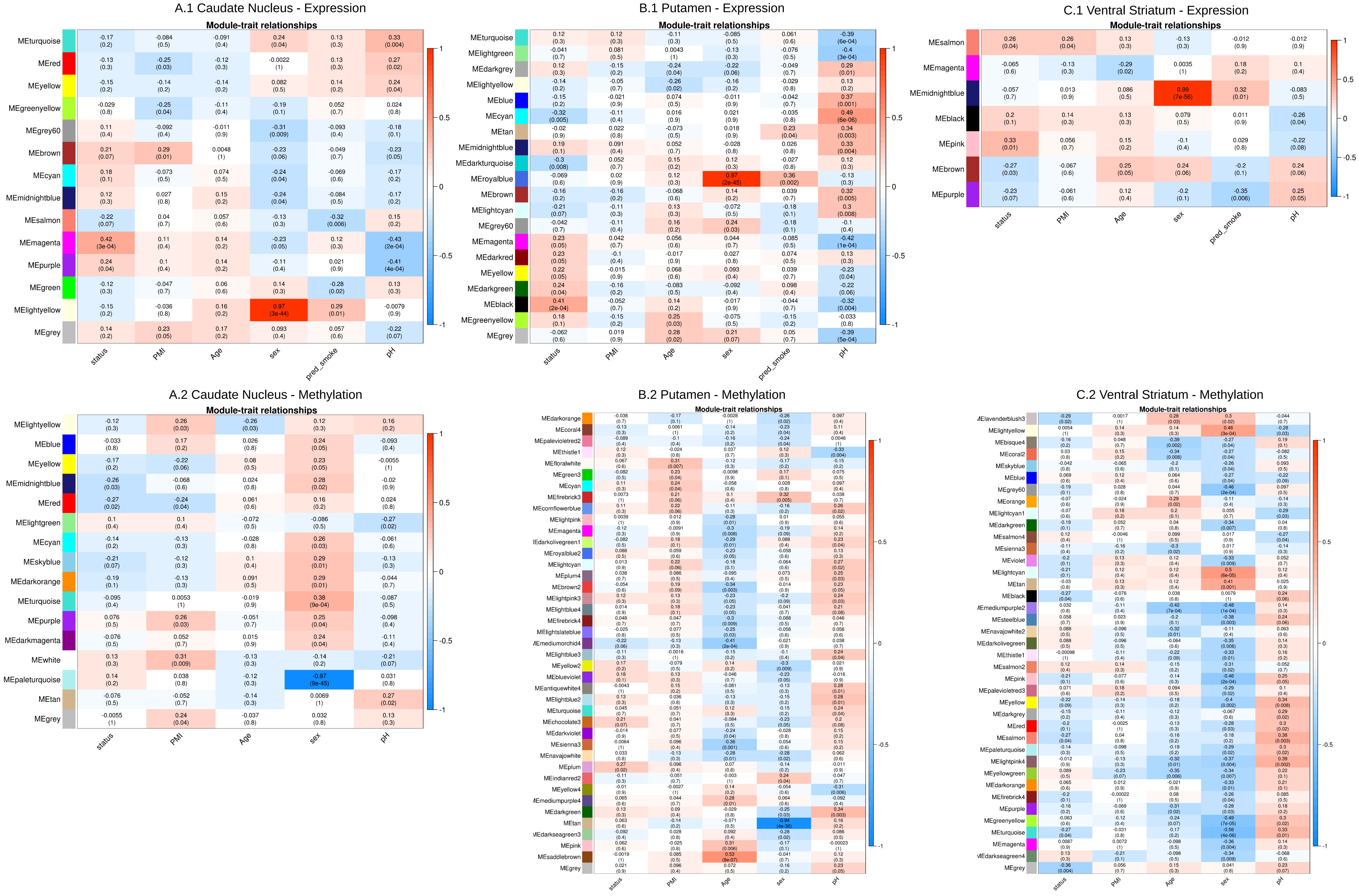

### Supplementary Figure 5

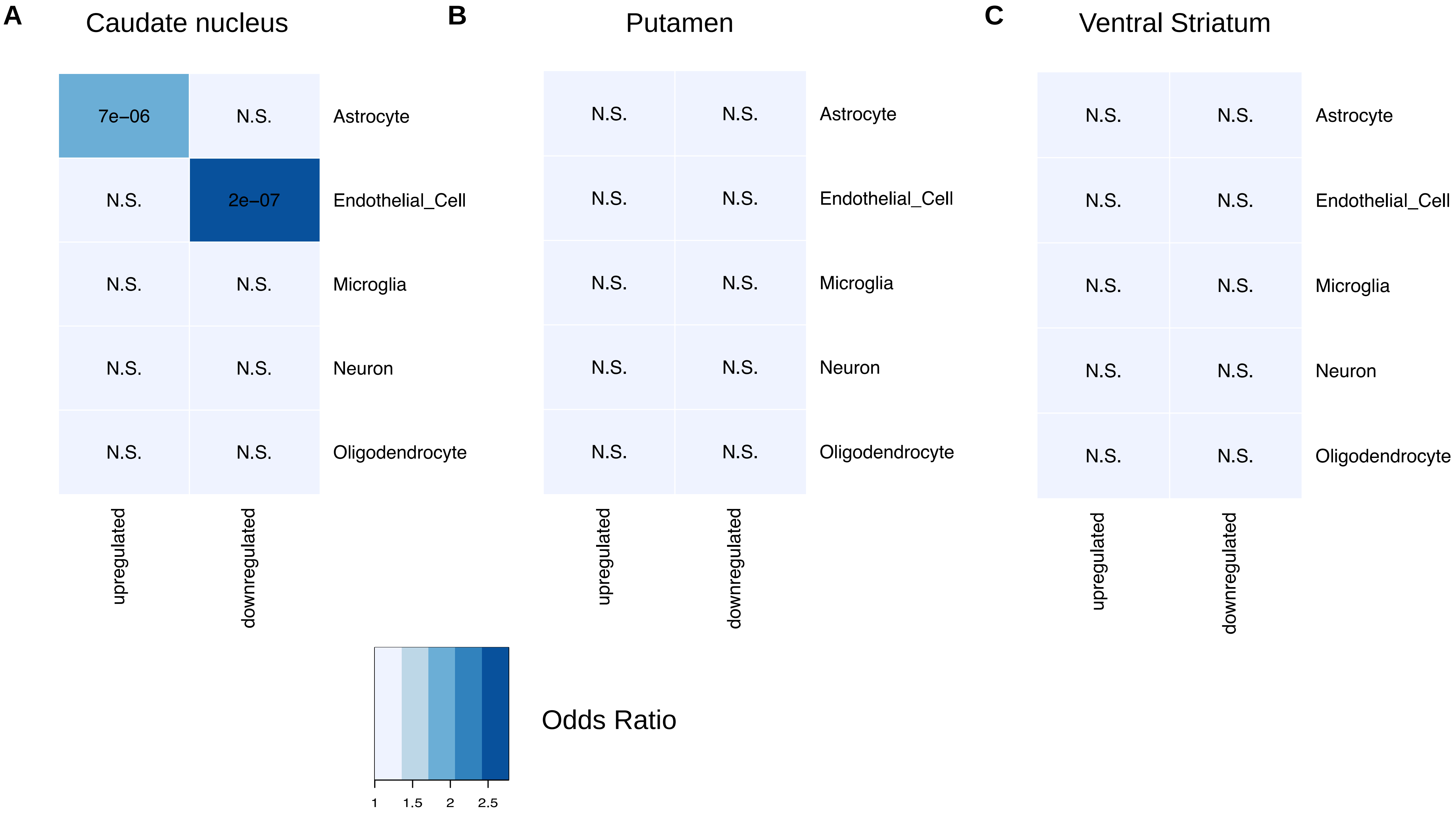

### Supplementary Figure 6

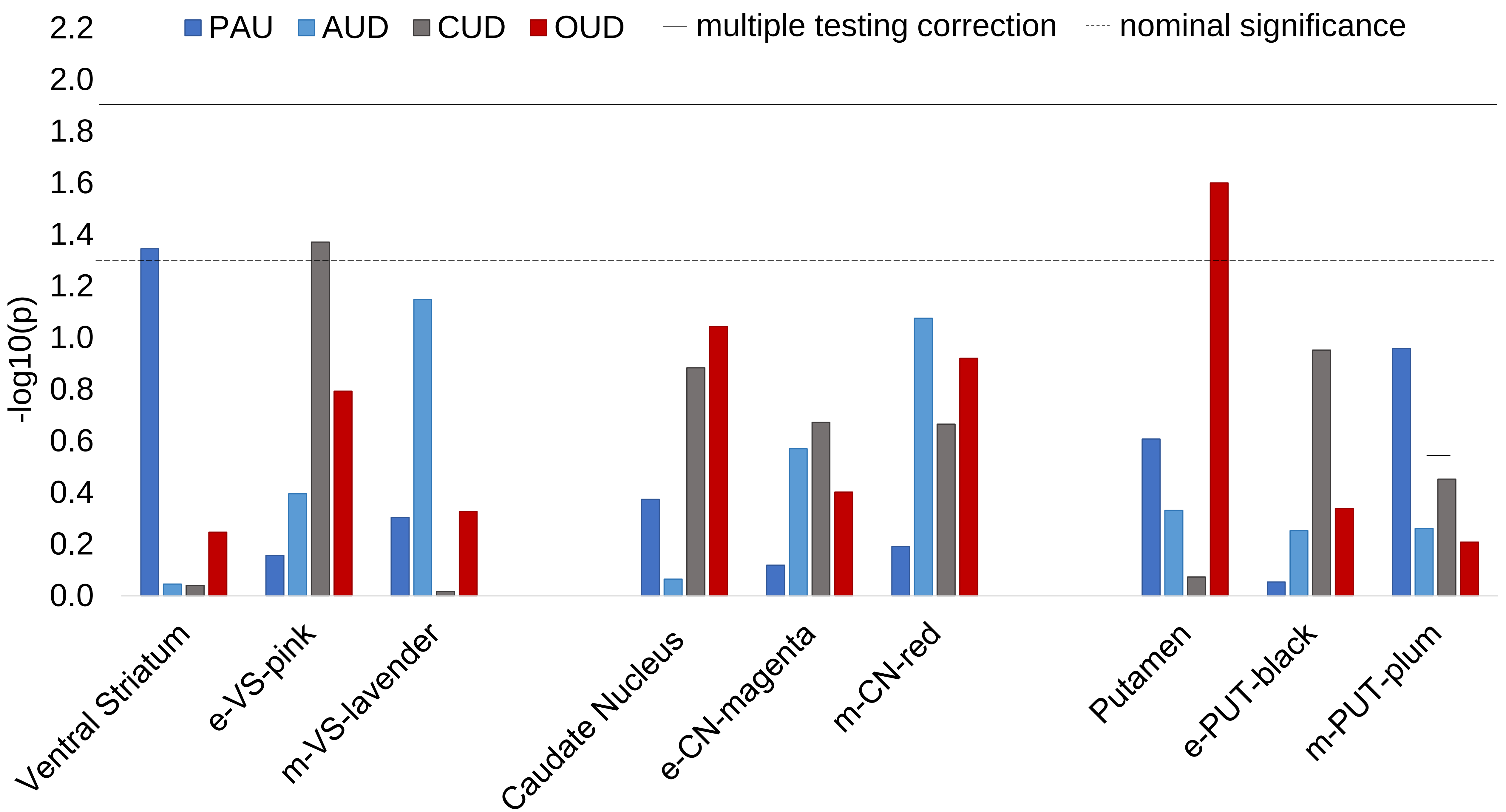
