## Supplementary Figure Legends for "Multi-Omics Signatures of Alcohol Use Disorder in the Dorsal and Ventral Striatum"

Figure Legends Supplementary Figures

**Supplementary Figure 1.** QQ-plots for the differential expression analysis in A) caudate nucleus, B) putamen and C) ventral striatum.

**Supplementary Figure 2.** Variance partition analysis of gene expression data from A) caudate nucleus, B) putamen and C) ventral striatum.

**Supplementary Figure 3.** Volcano plots of the differential expression analysis in A) caudate nucleus, B) putamen and C) ventral striatum. Red dots indicate significance at FDR<0.25, green dots a log2(foldchange) > 0.2.

**Supplementary Figure 4.** Heatmaps of the module trait relationships from WGCNA. A. disaplys data from caudate nucleus, B. from putamen and C. from ventral striatum. Methylation modules have the suffix 1 and expression modules suffix 2.

**Supplementary Figure 5.** Heatmaps of cell-type enrichment of up- and downregulated genes in AUD (FDR<0.25) for A) caudate nucleus, B) putamen and C) ventral striatum.

**Supplementary Figure 6.** Bar plots depicting the –log10 transformed p values of GWAS enrichment analysis for DE genes and the WGCNA expression and methylation modules showing the strongest association with AUD in all three brain regions. Color represents the GWAS: dark blue = alcohol use disorder (AUD), light blue = problematic alcohol use (PAU), grey = Cannabis use disorder (CUD) and red = opioid use disorder (OUD). Dotted line represents nominal significance, solid line depicts significance threshold after multiple testing correction.
